# Hierarchical Representation of Complex Intervention Sequences for Automated Subgroup Analysis in Critical Care Settings

**DOI:** 10.1101/2021.10.16.21265074

**Authors:** Amee Trivedi, William Ogallo, Girmaw Abebe Tadesse

**Author notes:** **Address for correspondence** Amee Trivedi.

## Abstract

Our understanding of the impact of interventions in critical care is limited by the lack of techniques that represent and analyze complex intervention spaces applied across heterogeneous patient populations. Existing work has mainly focused on selecting a few interventions and representing them as binary variables, resulting in oversimplification of intervention representation. The goal of this study is to find effective representations of sequential interventions to support intervention effect analysis. To this end, we have developed Hi-RISE (Hierarchical Representation of Intervention Sequences), an approach that transforms and clusters sequential interventions into a latent space, with the resulting clusters used for heterogenous treatment effect analysis. We apply this approach to the MIMIC III dataset and identified intervention clusters and corresponding subpopulations with peculiar odds of 28-day mortality. Our approach may lead to a better understanding of the subgroup-level effects of sequential interventions and improve targeted intervention planning in critical care settings.

## Introduction

Healthcare is characterized by multi-dimensional variations across patient populations and care interventions that differentially impact healthcare outcomes and costs [1]. Patient variations arise from differences in characteristics such as age groups, gender, socioeconomic status, and comorbidities, and have been shown to result in different manifestations of diseases and treatment effects across predefined subpopulations. Variations in interventions arise from differences in the type, number (*single* vs. *multiple)*, and longitudinal connectivity (*concurrent* vs. *sequential*) of interventions used in healthcare. In critical care settings, effective analysis of variations of care could improve clinical outcomes and decrease the rising costs of critical care[2,3].

The effective representation of interventions is crucial to discover their significant impacts across subpopulations. Particularly, this becomes more challenging when multiple interventions are involved that might have different degrees of longitudinal overlapping. Concurrent interventions refer to those sharing a temporal window when they are applied. On the other hand, sequential interventions don’t possess significant overlapping in their application windows temporally, and one is applied after the other. In addition, an intervention could be just applied once *(static)* or applied frequently *(dynamic)*. Thus, it becomes clear that to underline the impact of the intervention(s) on care outcome, it is crucial to have effective representations of interventions that are robust to their variations and complexity.

Existing research in subpopulation-based intervention effect analysis is confined to a priorly defined group of interest (e.g., for rural community compared to urban) and the intervention is limited to a single and static intervention with binary arms (1= intervention applied, 0= absence of intervention). The requirement of presupposing the group of subpopulations of interest limits the discovery of data-driven insights (that were not priorly understood by domain experts). On the other hand, the existing impact analyses involving single binary interventions tend to have the following limitations: 1) in cases of multiple interventions, simple binarization for each intervention lacks the capability to encode overlapping time-windows of their applications, i.e., concurrent impact. 2) in addition, binary representation could not encode detailed granularities of the intervention(s) applied, e.g., dosage amount, mode of administration, etc.; 3) existing intervention representation techniques also fail to encode long-term temporal dependency of sequential interventions, e.g., a late side-effect or a long-term correlation with another intervention, which might be overlooked by manual intervention selection. Moreover, analysis of sequential interventions needs to be flexible to work on different levels of intervention resolutions and to provide efficient visualization to communicate extracted insights.

In this paper, we aim to address the challenges associated with effective representation of sequential interventions and to extract insights on the effects of these interventions across subpopulations without the need of pre-supposing these subpopulations, i.e., *automatically identify the subpopulations with the highest impact by a sequence of interventions*. To overcome the problem of sequence learning and representation in a large space, we use heuristics derived from empirical analysis and the use of autoencoder-based clustering that helps to reduce the representation space without losing its intelligibility and aggregates multiple interventions based on their late-based similarity. To summarize, the main contributions of this paper are twofold. First, we present an approach to identify and represent sequential interventions from a large complex state space to a small latent variable state space. This helps in the representation of large numbers of interventions, learning correlations between multiple interventions, and accounting for the intervention order. Second, we present a pipeline to identify the intervention sequence and user subpopulation as anomalous patterns of care. We evaluate our approach using a real-world MIMIC-III dataset and present the findings.

## Methods

### Approach

Towards addressing the challenges associated with representing and analyzing the subgroup-level impact (heterogeneous effects) of complex intervention sequences, we propose a modular approach consisting of 4 main steps: *data cleaning and standardization, hierarchical representation of intervention sequences, dimensionality reduction and clustering*, and *automated subgroup analysis*.

### Data cleaning and standardization

The data cleaning and standardization module takes as input raw sequential intervention data captured in longitudinal electronic health records and provides a standardized representation of the care providers and interventions. It is responsible for operations such as dealing with missing values as well as removing redundant and irrelevant sequential intervention data. It is also used to standardize the sequential intervention data through operations such as recognition and resolution of different abbreviations for the same care provider type, event type, or intervention.

The module only considers the intervention events in electronic health records. For each patient, these events are ordered temporally such that the final output of this module is a clean standardized sequential intervention dataset which is fed to the next module in the pipeline.

### Hierarchical representation of intervention sequences

The main objective of the Hierarchical representation of intervention sequences (Hi-RISE) module is to compactly represent preprocessed sequential interventions at multiple intervention granularities. Each clinic visit by a patient results in a sequence of care provider encounters. Each care provider has a specific set of duties and hence generates a set of events such as patient registration, vitals measurements, lab tests, medication administration, a recommendation for other tests, etc.

The granularity of the intervention representation is governed by the application or intervention space usage. Some applications might need more fine-grained intervention space representations that incorporate low-level intervention details such as dosage administered, mode of administration, and frequency of dosage others might only need care provider encounter sequence. To compactly represent events generated during patient encounters with care providers in increasing order of granularity for interventions we use a hierarchical representation of the space through Hi-RISE. Currently, Hi-RISE is designed to consists of 3 main levels to maintain its simplicity. The levels are labeled as “L0”, “L1”, and “L2” as illustrated in Figure 1.

**Figure 1.**
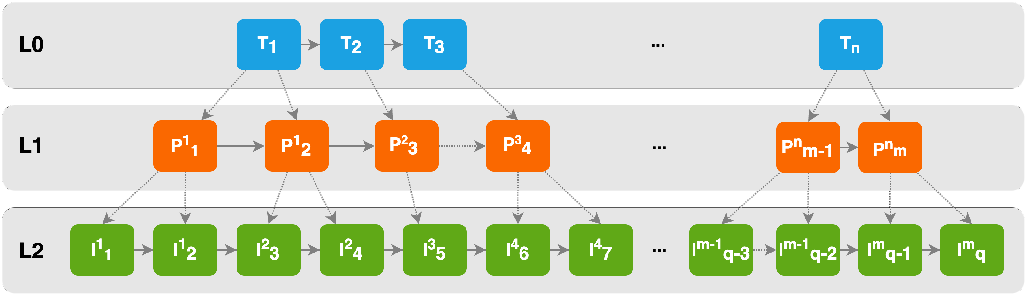
Illustration of Hi-RISE (**Hi**erarchical **R**epresentation of **I**ntervention **Se**quences). L0 represents the sequence of provider types (T_1_…T_n_), L1 represents the sequences of actual provider encounters 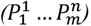, and L2 represents sequences of interventions 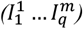.

L0 is the coarsest level of representation in Hi-RISE. Here, we aggregate all contiguous interventions from the same type of care providers as a supercluster representation of care provider type encounters. The entire sequence of care provider encounters for a patient will be represented as a supercluster sequence of care providers based on the care provider type. An example of a sequence at L0 is *nurses → doctors → nurses → lab technicians → nurses → doctors*.

L1 increases the granularity of L0 by representing the actual sequence of care provider encounters in a patient’s care journey. An example of such an L1 sequence is *nurse A → doctor A → nurse B → nurse C → senior nurse A → lab technician A → nurse B → doctor B*.

L2 represents actual interventions during individual care provider encounters. An example of an L2 sequence is *interventions [1,2] (nurse A) → interventions [3,4] (doctor A) → intervention 5 (nurse B) → interventions [6,7,8] (nurse B) → intervention 9 (doctor A)*. Each caregiver can trigger a series of events in any order. However, rather than imposing fixed temporal orders on the events generated by care providers during each encounter, we use a “bag-of-interventions” (similar to “bag-of-words”[9]) representation of events per care provider. The bag-of-interventions representation ensures that the intervention or events noted per care provider per encounter do not have any order to them. Each intervention is a one-hot-encoded representation, and each bag-of-interventions is computed as the sum of all the one-hot-encoded representations of the interventions administered by a provider at a point in time. The reasoning behind using the bag-of-interventions approach is that the medicines registered by a care provider at a given point in time encounter might not necessarily need to follow an order.

Mathematically, Hi-RISE can be formulated is as follows:

If E is a patient that visits a care provider resulting in a sequence of care provider encounters represented by *P* = {*P*_1_, *P*_2_, …, *P*_*n* − 1_, *P*_*n*_}, and a sequence of interventions, *I* = {*I*_1_, *I*_2_, …, *I*_*n*− 1_, *I*_*n*_},

We define:

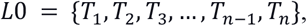

where *T*_*i*_ *≠ T*_*i* + 1_ *for* 1 ≤ *i* ≤ *n*

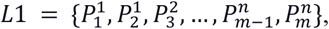

where *∀ i, j ProviderType* 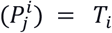 *for* 1 ≤ *i* ≤ *n*

Intervention sequence = 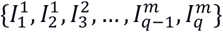,

where *∀ j, k CareProvideer* 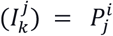 *for* 1 *≤ j ≤ m*

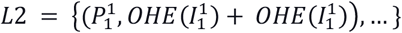

and *CareProvideer*(*I*_*a*_) = *P*_*b*_, where *P*_*b*_, is the care provider that administered the intervention *I*_*a*_;

*ProvideerType*(*P*_*b*_) = *T*_*c*_ where *T*_*c*_ is the care provider type of provider *P*_*b*_.

*OHE*(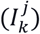) is the One hot encoding of the Intervention *I*_*k*_.

The multi-level outputs of the Hi-RISE module are critical in simplifying care provider interventions observed in longitudinal patient data. However, these outputs are still complex and not readily applicable to current approaches needed by sub-group-level intervention impact analysis.

### Dimensionality reduction and clustering

The outputs of the Hi-RISE module serve as inputs to the dimensionality reduction and clustering module. The function of this module is to logically identify groups of similar sequences across multiple patients per each Hi-RISE level. The module consists of an autoencoder[8] followed by K-Means clustering[7]. An autoencoder is an unsupervised artificial neural network that learns efficient data encoding to reduce the dimensionality and produce a compact latent space representation of the data aka sequential interventions in our case. The output from the autoencoder is fed to a K-Means clustering submodule. K-Means clustering is a vector quantization technique to partition the autoencoder latent space representations of the sequence interventions into k partitions. The objective of K-Means clustering is to minimize the intra-cluster variances with squared Euclidean distance as the metric. K-Means creates clusters of the compact intervention sequences from the autoencoder based on representation similarity.

For each transformed intervention sequence cluster, C, we create a binary intervention, Z such that for each patient, Z=1 if the patient’s sequence of interventions can be mapped to C, and Z=0 otherwise. Next, we model the propensity score (*p*_*s*_) as the probability of the intervention conditioned on observed baseline covariates, i.e., *p*_*s*_(*X*) = *p*(*Z* = 1|*X*), where X is the set of covariates.

### Automated subgroup analysis

The automated subgroup analysis module is to evaluate the heterogeneous treatment effects of the transformed intervention sequences to identify the subgroups of patients whose outcomes are most/least impacted by the interventions. In addition to the transformed intervention sequence clusters, this module takes as input covariates (e.g., age, gender, socioeconomic status, comorbidities, etc.) and an outcome of interest (e.g., length of stay, readmission, etc.). These inputs are analyzed using an algorithm that combines inverse propensity of treatment weighting (IPTW)[1] followed by automated subgroup analysis via subset scanning[6].

IPTW eliminates bias due to observable differences between the treated and non-treated/comparison groups. Note that the IPTW would not be necessary in the case of a randomized study. The automated subgroup analysis is used to discover the subgroups of patients whose outcomes are most impacted by interventions in the unbiased dataset. We frame this as a search problem and build our approach upon the Bias Scan algorithm[10]. We use the average treatment effect on the treated (ATT) weights and quantify the anomalousness of a subpopulation, *S*, as *E*[*w*_*i*_*Y*_*i*_(1)|*X*_*i*_ ∈ *S*] < *E*[*w*_*i*_*Y*_*i*_(0)] for subjects with lower than expected outcomes and *E*[*w*_*i*_*Y*_*i*_(1)|*X*_*i*_ ∈ *S*] > *E*[*w*_*i*_*Y*_*i*_(0)] for subjects with high than expected outcomes. These subgroups are characterized by a subset of covariates that are identified to maximize the outcome divergence score from the expected.

### Experiment with MIMIC III

#### Dataset and Cohort

To evaluate our approach, we conducted experiments on the MIMIC-III (Medical Information Mart for Intensive Care), a publicly available critical care database[4]. The study population consisted of 46,520 patients with 57,786 hospital admissions and 61,532 Intensive Care Unit (ICU) stays. Our study cohort included adult patients (16 years or older) admitted to the ICU for the first time. We excluded patients with a length of stay less than 1 day, hospital readmissions, surgical cases, or having chart events. The final cohort consisted of 18,761 patients.

#### Covariates, Outcome, and Interventions

We conducted feature extraction and preprocessing to generate a final dataset that consisted of a binary outcome (mortality within 28 days), and 13 discrete covariates as follows: age (65 or older, <65), gender (female, male), ethnicity (Black/Africa, Caucasian, other, unknown), marital status (married/partner, single, divorced/separated/widowed, unknown), insurance (private/self-pay, Medicare/Medicaid), arrhythmia (no, yes), chronic pulmonary disease (no, yes), congestive heart failure (no, yes), diabetes (no, yes), Angus sepsis (no, yes), urine output (high, low, normal), ventilated (no, yes). Additionally, for all patients in the cohort, we extracted and transformed the patients’ intervention sequences and subsequently generated 78 binary interventions representing patients with similar intervention sequences. Finally, we filtered out interventions with fewer than 20% of the observations and ended up with 10 interventions that were analyzed further.

#### Subgroup analysis

For each of the 10 binary interventions, we trained a logistic regression model and used it to predict the likelihood of a patient having the intervention given his/her covariates. We opted for the logistic regression approach due to its simplicity and interpretability. The AUC of the models ranged between 56% and 72%. Next, for each intervention, we computed the ATT weights using the corresponding propensity score model and used these to compute the expected weight mean of the outcome, which served as the counterfactual outcome for each subject. Finally, we used our automated subgroup analysis approach to search over the treated subjects in the final dataset to identify the treatments and corresponding subpopulations that showed the most evidence of having outcomes rates that differed the most from the weighted mean of the expected outcome rate. For each intervention, this search identified the subgroup having the highest odds of 28-day mortality (positive direction) and the subgroup having the lowest odds of 28-day mortality (negative direction).

## Results

### Representation descriptive statistics

Post preprocessing and data cleaning, we found that on average, there are 5442 events generated per hospitalized patient. A patient-careprovider Sankey flow graph of the top 31 interactions between care provider pairs is shown in Figure 2.

**Figure 2.**
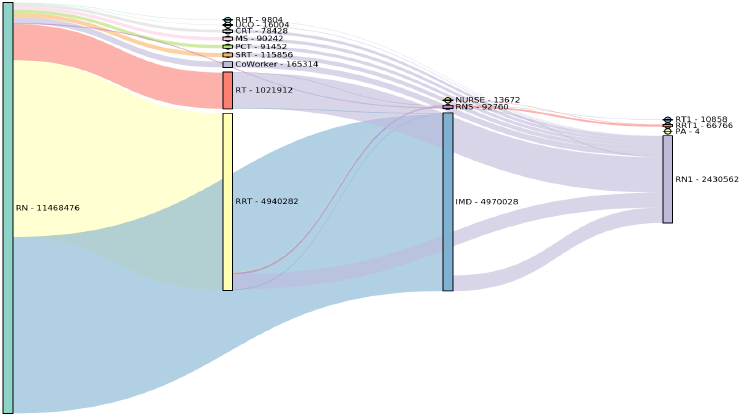
Sankey plot of top 31 provider-patient interactions pairs.

The top 2 common patient intervention sequence interactions are with Registered Nurse (RN) followed by Rapid Response Team (RRT) or Registered Nurse (RN) followed by a Medical Doctor (IMD). This helps us realize that few subsequences are extremely common but not all subsequences start with an RN followed by an RRT or MD. Learning the relationships between these infrequent, out-of-order, or non-encounters with the care providers could result in a favorable or non-favorable outcome based on the patient attributes.

The total number of patient care provider items in our dataset is large so we divide the items as *monitoring events* or *interventions* and then use Hi-RISE to compactly represent the intervention sequences. Figure 3 shows the histogram of the number of sequence lengths.

**Figure 3.**
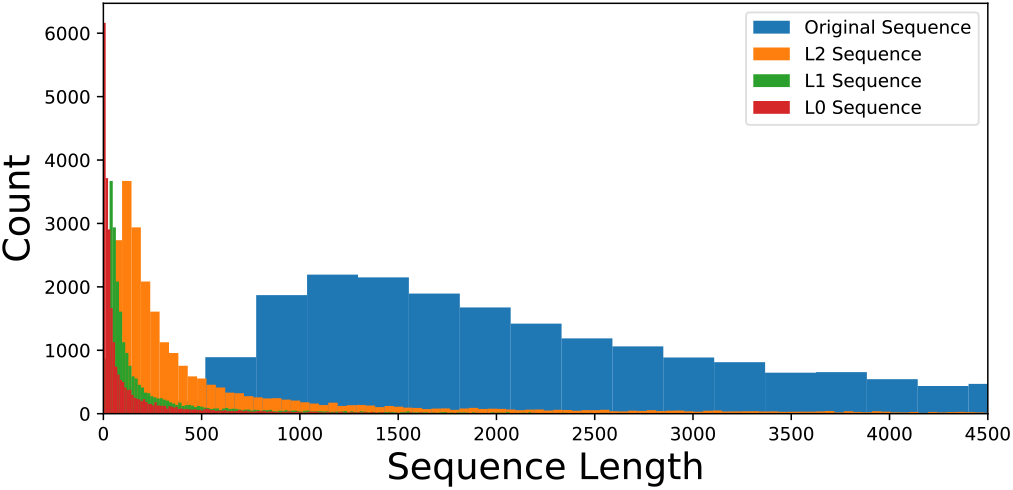
Histograms of sequence lengths of the original intervention sequence and Hi-RISE’s L0, L1, and L2 sequence representations.

The mean number of items per intervention sequence in the dataset is 2533. Hi-RISE reduces the intervention sequence length to 39 for L0, 90 for L1, and 270 for L2 as shown in figure X. Please note that the order of care provider encounter is preserved at each hierarchical level.

### Dimensionality reduction and clustering

As shown above, the dimensionality of Hi-RISE representation is still high. We use an autoencoder for further dimensionality reduction. The hyperparameters of the autoencoder are set as follows: Adam optimizer, binary cross-entropy loss, 256 training epochs with a batch size of 256, Relu and Sigmoid activation functions for the encoder and decoder, respectively. We have also experimented the autoencoder with different latent-space dimensions. For clustering the compressed latent space representation of the intervention sequence, we select the number of clusters based on the k-means inertia score, which is a measure of the internal cluster coherence. It is measured by the sum of squared distances of the points to the nearest cluster center. The inertia value plot and clusterings are shown in Figure 4.

**Figure 4.**
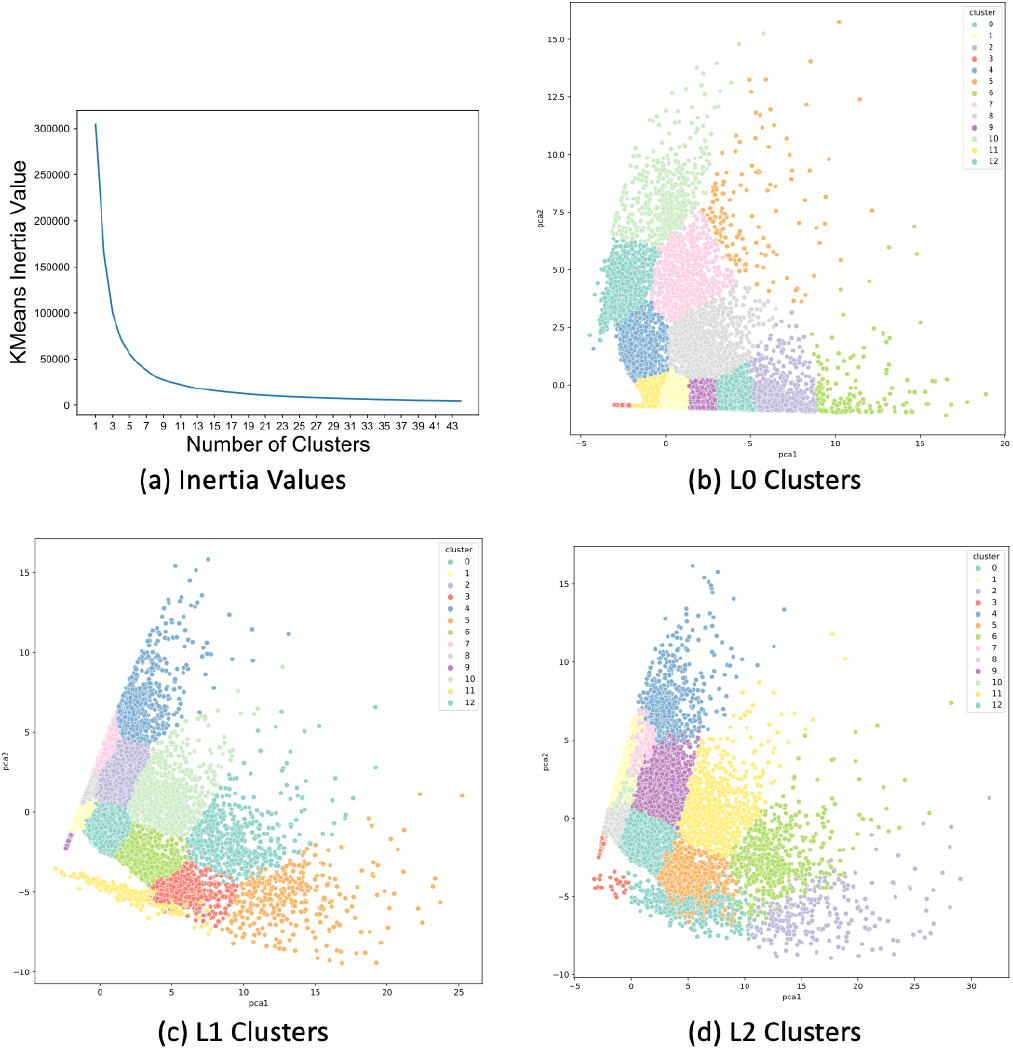
K-means inertia scores and clustering

### Characteristics of the anomalous subgroups identified

Table 1 shows the anomalous pattern detection results for the different intervention clusters extracted from MIMIC III. Each row represents a single cluster of sequential interventions generated from a particular Hi-RISE level and a specific autoencoder dimension. In the negative direction, we note that the search algorithm identified subpopulations of individuals in each cluster where the odds of 28-day mortality in the subpopulation were much lower than the odds in the overall population. Similarly, in the positive direction, the algorithm identified subpopulations in which the odds of 28-day mortality were up to 2.8 times higher in the subpopulations, whereas the odds of the outcome were lower in the overall population.

**Table 1.** Anomalous subgroup analysis results

| Dir | L | D | C | P | Anomalous Subpop |  | Odds Ratio |  |
| --- | --- | --- | --- | --- | --- | --- | --- | --- |
|  |  |  |  |  | Size | Score | Pop | Subpop |
| Neg | 0 | 9 | 3 | 0.27 | 1974 (39%) | 151 | 0.8 | 0.2 |
|  | 0 | 48 | 3 | 0.28 | 2032 (38%) | 152.2 | 0.8 | 0.2 |
|  | 1 | 9 | 6 | 0.22 | 1594 (39%) | 124.7 | 0.8 | 0.2 |
|  | 1 | 48 | 0 | 0.29 | 2119 (38%) | 157.7 | 0.8 | 0.2 |
|  | 2 | 9 | 6 | 0.22 | 1594 (39%) | 124.7 | 0.8 | 0.2 |
|  | 2 | 48 | 0 | 0.29 | 2119 (38%) | 157.7 | 0.8 | 0.2 |
| Pos | 0 | 9 | 3 | 0.27 | 621 (12%) | 66.1 | 0.8 | 2.7 |
|  | 0 | 48 | 3 | 0.28 | 644 (12%) | 73.1 | 0.8 | 2.8 |
|  | 1 | 9 | 6 | 0.22 | 513 (12%) | 50.8 | 0.8 | 2.7 |
|  | 1 | 48 | 0 | 0.29 | 670 (12%) | 73.8 | 0.8 | 2.8 |
|  | 2 | 9 | 6 | 0.22 | 513 (12%) | 50.8 | 0.8 | 2.7 |
|  | 2 | 48 | 0 | 0.29 | 670 (12%) | 73.8 | 0.8 | 2.8 |

Dir: Direction of Scanning, L: Hi-RISE level, D: Autoencoder dimension, C: cluster, P: proportion in cluster, Pop: Population, Subpop: Subpopulation

## Discussion

In this work, we proposed Hi-RISE, a simple heuristic approach to represent complex sequences of care provider interventions at various granularities. We employ an autoencoder to further reduce the dimensionality of Hi-RISE sequences and then cluster the encoder representations. These clustered sequential intervention sequences along with patient covariates are fed to an automated subgroup analysis algorithm to identify subpopulations with the most positive or negative outcome for a sequential intervention. We demonstrate the application of our approach to the MIMIC III dataset.

The key insight is that even small variations in intervention sequences can change the outcome of patient care, so we should consider interventions as an ordered sequence and not a set of few hand-picked variables. Additionally, the task of finding similarity between intervention sequences of a large number of patients is a computationally expensive and resource-heavy task and we overcome this limitation in our computationally fast approach that is resource sustainable, and robust to noise also. Our current approach of using an autoencoder is not easily interpretable design and as a follow-up to this work, we would like to research interpretable methods for dimensionality reduction while paying attention to the ordering.

Researchers have previously investigated the heterogeneous treatment effects for binary outcomes[1] and interventions[5]. To the best of our knowledge, this work is one of the first to specifically investigate how complex heterogeneous treatment effects of sequential interventions could be studied in a simplified manner. However, this work is only preliminary and the approach is limited to generating hypotheses about specific populations most impacted by certain intervention sequences. Furthermore, this paper does not report on the individual characteristics of the anomalous subgroups. Additional analyses and characterizations of these anomalous subgroups are warranted.

## Conclusions

This study showed that ordering of intervention events is important and that it is possible to succinctly represent the intervention space in smaller dimensions to support heterogeneous treatment effect analysis. Our next steps are to make our approach more interpretable while adhering to resource sustainability. We also plan to use our approach in other domains such as COVID-19 data analysis, for which different sequences of interventions might be applied across space and time.

## Data Availability

MIMIC-III Public Open Dataset

## Notes

### Competing Interest Statement

The authors have declared no competing interest.

### Funding Statement

This study did not receive any funding

